# Long-term monitoring of SARS-CoV-2 in wastewater of the Frankfurt metropolitan area in Southern Germany

**DOI:** 10.1101/2020.10.26.20215020

**Authors:** Shelesh Agrawal, Laura Orschler, Susanne Lackner

## Abstract

Wastewater-based epidemiology (WBE) is a great approach that enables us to comprehensively monitor the community to determine the scale and dynamics of infections in a city, particularly in metropolitan cities with a high population density. Therefore, we monitored the time course of the SARS-CoV-2 RNA concentration in raw sewage in the Frankfurt metropolitan area, the European financial center. To determine the SARS-CoV-2 concentration in sewage, we continuously collected samples from two wastewater treatment plant (WWTP) influents (Niederrad and Sindlingen) serving the Frankfurt metropolitan area and performed RT-qPCR analysis targeting three genes (N gene, S gene, and ORF1ab gene). In August, a resurgence in the SARS-CoV-2 RNA load was observed, reaching 3 x 10^13^ copies/day, which represents similar levels compared to April with approx. 2 x 10^14^ copies/day. This corresponds to an also continuous increase again in COVID-19 cases in Frankfurt since August, with an average of 28.6 incidences, compared to 28.7 incidences in April. Different temporal dynamics were observed between different sampling points, indicating local dynamics in COVID-19 cases within the Frankfurt metropolitan area. The SARS-CoV-2 load to the WWTP Niederrad ranged from approx. 4 x 10^11^ to 1 x 10^15^ copies/day, the load to the WWTP Sindlingen from approx. 1 x 10^11^ to 2 x 10^14^ copies/day, which resulted in a preceding increase in these loading in July ahead of the weekly averaged incidences. The study shows that WBE has the potential as early warning system for SARS-CoV-2 infections and as monitoring system to identify global hotspots of COVID-19.

## Introduction

The ongoing pandemic of the coronavirus disease 2019 (COVID-19) is a public health emergency of global concern and is expressed by symptoms like fever, myalgia, fatigue, dry cough. The disease, caused by severe acute respiratory syndrome coronavirus 2 (SARS-CoV-2), emerged in China in December 2019 and the World Health Organization (WHO) declared it as pandemic on March 11^th^, 2020 due to its worldwide spread ^1^. The disease has now been reported in over 213 countries with more than 30 million confirmed cases. The pandemic has caused nationwide lockdowns in many countries and contact restrictions as a prevention for the spread of the disease ^2^.

In the last couple of month, wastewater epidemiology (WBE) has emerged as a promising approach for early warning of disease outbreaks and providing information for the public especially if patients are asymptotic. Recent studies confirmed the detection of SARS-CoV-2 in feces and urine from positive tested patients ^3,4^, which implies, that SARS-CoV-2 is present in the influent of wastewater treatment plants (WWTPs) ^5^.

The potential advantage of environmental surveillance in WBE is to enable prediction of the overall status of a given catchment area with much less effort than clinical surveillance. WBE can give an insight of the outbreak situation in the entire catchment area by testing wastewater sample over time. In contrast, clinical surveillance requires more time and cost for sample collection and testing. An additional big advantage of WBE is also capturing people with asymptomatic and pre-symptomatic infections, who may not be included in clinical surveillance. Several studies have already proved that wastewater monitoring can detect outbreaks of norovirus and poliovirus earlier than clinical surveys ^6–9^.

Preliminary studies have reported the detection of SARS-CoV-2 RNA in wastewater in the Netherlands ^10^, USA ^11^, Australia ^12^ and Italy ^13^. One of the first studies based on surveillance of COVID-19 in wastewater was performed in Australia and SARS-CoV-2 was detected in two samples within a six day period of the same WWTP with both qPCR and sequencing ^12^. In the Netherlands, researchers tested sewage of six cities and the airport for SARS-CoV-2, targeting either the nucleocapsid (N) gene or the envelope (E) gene ^10^. The results showed that the sewage samples were tested positive for the N gene in March 2020. Whereas in Italy, a research group studied twelve influent sewage samples from the WWTPs in Milan and Rome between February and April with the result that 6 out of 12 samples were positive ^13^.

In this study, the goal was to establish a WBE surveillance system for SARS-CoV-2 in a metropolitan area in Southern Germany (Frankfurt am Main) and to use this data as warning system in the future. WWTP data can add valuable information and thus aid decision making on further public and societal restrictions or easings with increasing or decreasing virus concentration.

## Material and Methods

### Sewage Samples

24h flow-proportional samples were collected between April 2020 and August 2020 at three different sampling points from two wastewater treatment plants (WWTP), located in Frankfurt am Main, Germany. The WWTP Niederrad/Griesheim is designed for a population equivalent (PE) of 1.350.000, the WWTP Sindlingen for 470.000 PE. These two WWTPs receive the sewage of approximately 1.200.000 people, the remaining PE can be allocated to commercial and industrial discharges. Table 1 also includes the 85 % percentiles of the influent volumes. Daily readings of the respective flow volume’s were used to calculated SARS-CoV-2 loadings. In this study, we processed 44 samples, which include 17 samples from Niederrad sampling point, 14 from Sindlingen sampling point, and 13 samples from Griesheim sampling point.

**Table 1:** Overview of the studied WWTPs in the city of Frankfurt am Main with the 85 % percentile influent volume per day for the period 15.06. – 31.08.2020 and population equivalents (PE).

| Location | PE | 85 % percentile influent volume<br>[m<sup>3</sup>/d] |
| --- | --- | --- |
| Frankfurt am Main, Niederrad/Griesheim | 1.350.000 | 121.875 / 152.030 |
| Frankfurt am Main, Sindlingen | 470.000 | 64.085 |

**Table 2:** Information about dyes corresponding to each target gene.

| Gene | Dye | Quencher |
| --- | --- | --- |
| ORF1ab | FAM | QSY |
| N Protein | VIC | QSY |
| S Protein | ABY | QSY |
| MS2<br>(Internal Positive Control) | JUN | QSY |

**Table 3:**
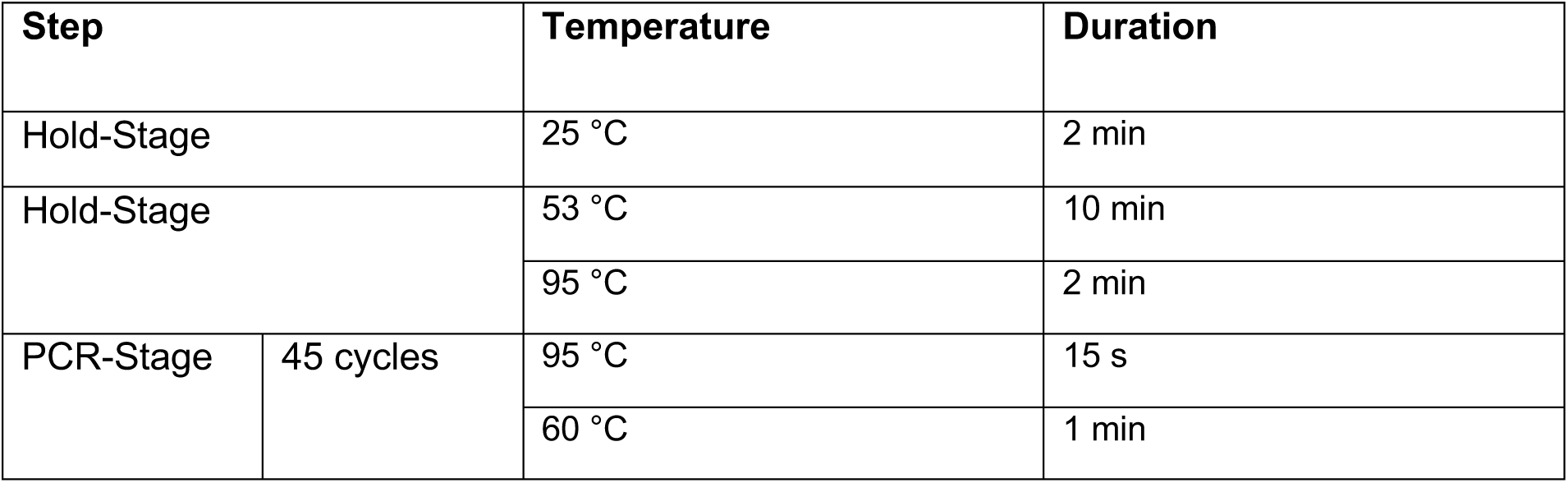
PCR protocol

For each sampling location, one liter of the untreated wastewater was spiked with 200 µL MS2 phage (Thermo Fisher Scientific) and filtered through a 0.45 µm electronegative membrane filter. The filters were divided and stored at minus 80°C prior to further downstream analysis.

RNA was extracted from the filter samples using the Fast RNA Blue Kit (MP Biomedicals) according to the manufacturer’s protocol and eluted with 100 µL of RNase free buffer. This RNA was used as a template for RT-qPCR. The concentration was measured using a Qubit 3.0 Fluorometer (Thermo Fisher Scientific).

### qPCR analysis

The RNA was analyzed using the Taq Path COVID-19 RT-PCR Kit (Thermo Fisher Scientific) with a Quant Studio 3 Thermal Cycler. This kit includes primer pairs targeting the N-, S- and ORF1ab genes were used as a multiplex assay. Details about the kit are provided in the supplementary information. Each qPCR run was performed in triplicates with 50 µL volume, with 12.5 µL TaqPath™ 1-Step Multiplex Master Mix (4X),2.5 µL COVID-19 Real Time PCR Assay Multiplex and 25 µL nuclease free water. To the reaction mix, 10 µL of purified and extracted viral RNA were added. Thermal profiles are provided in the supplementary information (Table 1). Reactions were considered positive if the cycle threshold was below 40 cycles. In each qPCR run, multiple SARS-CoV-2 RNA positive control, MS2 phage control (to determine the RNA recovery efficiency) of different known concentration and negative control were included, further details are provided in the supplementary information. The RT-qPCR abundance data were analyzed in R, using ggplot2 (v0.9.3.1), further details about the PCR efficiencies, threshold and baseline setting are provided in the supplementary information.

## Results and Discussion

### Epidemiological surveillance

We investigated the largest metropolitan region in southern Germany (Frankfurt am Main) using three separate sampling points in the catchment of the two large WWTPs (Table 1) and this data was compared to the COVID-19 cases in the area.

Epidemiological data on COVID-19 in the studied area was retrieved from the publicly available repository of the Robert Koch Institute (https://survstat.rki.de/Default.aspx). Figure 1 shows the positive COVID-19 cases in Germany compared to the positive cases of the city of Frankfurt am Main as reported weekly. The black line indicates the numbers in Germany with a high peak in March 2020 and the subsequent decline after the contact restrictions were imposed on the 22^nd^ of March. The blue line represents the COVID-19 cases of the city of Frankfurt am Main with 150 to 200 cases per week during the first peak in March and an increase of COVID-19 cases end of July. Frankfurt did not have a severe outbreak in spring with incidences not higher than 36 per 100.000 people the first week of April. The grey bar symbolizes the summer vacation of all schools in Hesse starting on the 06^th^ of July until the 14^th^ of August and marks a time period with increased travelling within Germany and Europe. This resulted in similarly or even slightly higher numbers of COVID-19 cases in the middle of August compared to April, whereas the overall trend for Germany only saw a slight increase in cases during that time. Additionally, it is necessary to mention, that during July and August the German government started free-of-cost testing for people returning from other countries as well as risk regions to control returnees from holiday. This may have also led to an increase in the reported positive COVID-19 case in the summer months.

**Figure 1:**
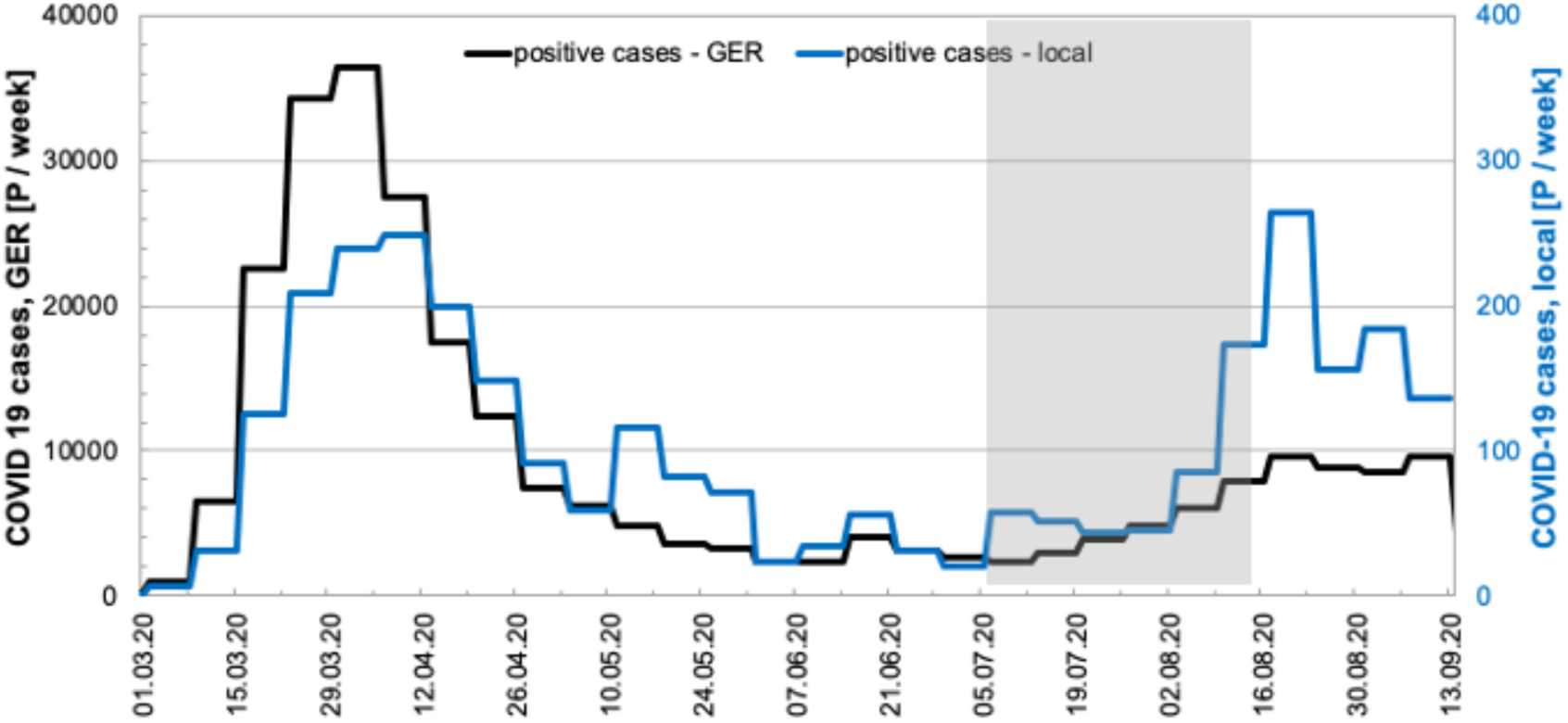
Graphical representation of the epidemiological data of COVID-19 cases in Germany (black line) and the city of Frankfurt am Main (blue line). The grey area indicates the time of the summer school vacation in the State of Hesse.

### Comparison of the results from April and August

The highest numbers of COVID-19 cases in the city of Frankfurt am Main occurred in April and August 2020, with 28 cases per 100.000 people as three-week average. In contrast, the trend throughout Germany showed a much higher first peak in April with 44 cases per 100.000 people at the maximum and only 10 in August. Therefore, we used the COVID-19 cases and the corresponding SARS-CoV-2 loads to the WWTPs both in April and August to compare both time points of the pandemic (Figure 2A). Our results revealed, that the load of SARS-CoV-2 virus in the wastewater samples was only slightly higher in April with a maximum of 1.4 x 10^15^ copies/d compared to a maximum of 5.37 x 10^14^ copies/d in August for individual influent wastewater samples. Additionally, we evaluated the correlation between the incidences reported against the sum of the SARS-CoV-2 load measured in all the samples (Figure 2B). It is clearly visible that with the increase in incidences, a corresponding increase in the viral load was observed as also shown in the Spearman’s rank correlation coefficient rs of 0.7464 (p (2-tailed) = 0.00217). However, there is a certain scattering in the data, which might be due to the variance in the measured viral load for different sampling points, as previously reported (Medema *et al*., 2020; Westhaus *et al*., 2020).

**Figure 2:**
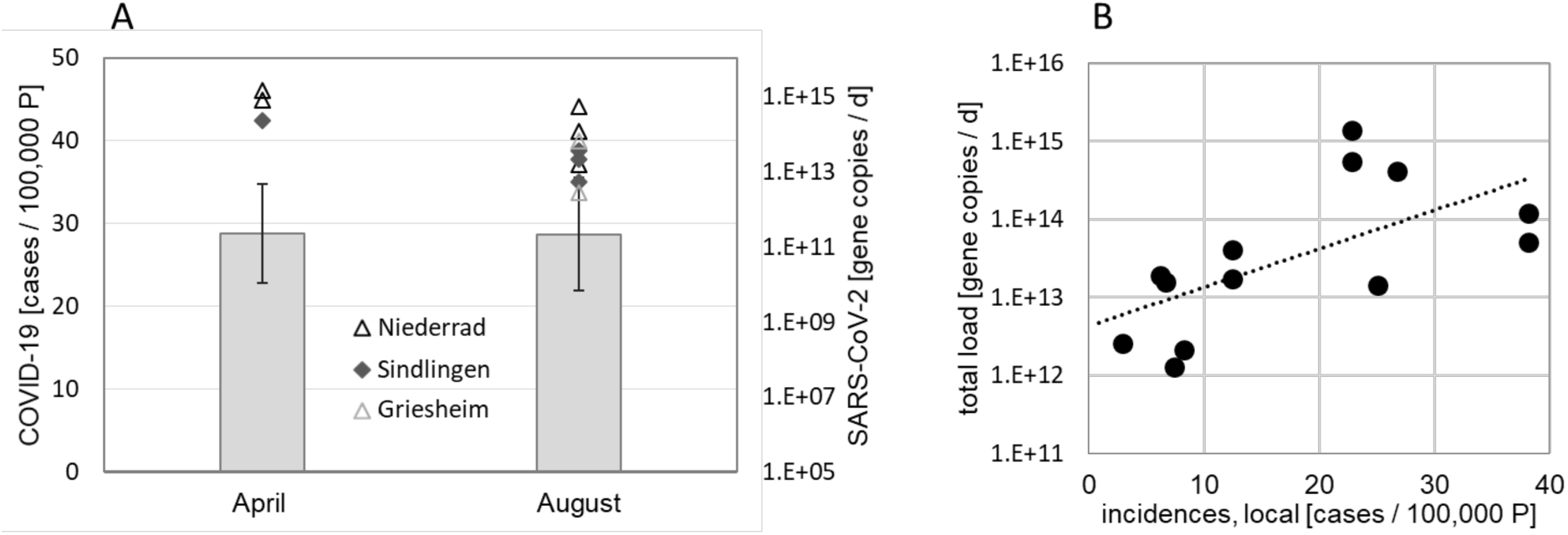
A: Comparison of the results from April and August 2020, bars represent a three week average around the maximum number of cases per 100.000 persons (P), symbols refer to the respective SARS-CoV-2 loads; B: Correlation between the total load of SARS-CoV-2 gene copies (sum of the loads to both WWTPs) and incidences in Frankfurt am Main (local)

### SARS-CoV-2 in Sewage Samples

The sum of the SARS-CoV-2 mean loads based on the concentrations of the three different target genes (N-, S- and ORF1ab gene) in the sewage samples from two WWTPs was compared to the reported positive tested COVID-19 cases in Frankfurt am Main (Figure 3). In the study period we observed that the SARS-CoV-2 loads ranged between 1.29 x 10^12^ copies/d on the 06.07.2020 as lowest and 1.63 x 10^15^ copies/d on the 21.04.2020 as highest. In June and July, the incidences were less than ten COVID-cases per 100.000 persons per week and the load ranged between 1.29 x 10^12^ and 1.91 x 10^13^ copies/d. The comparison with the COVID-19 cases revealed that the increase in the SARS-CoV-2 load clearly preceded the reported cases with the first step-increase in the middle of July 2020.

**Figure 3:**
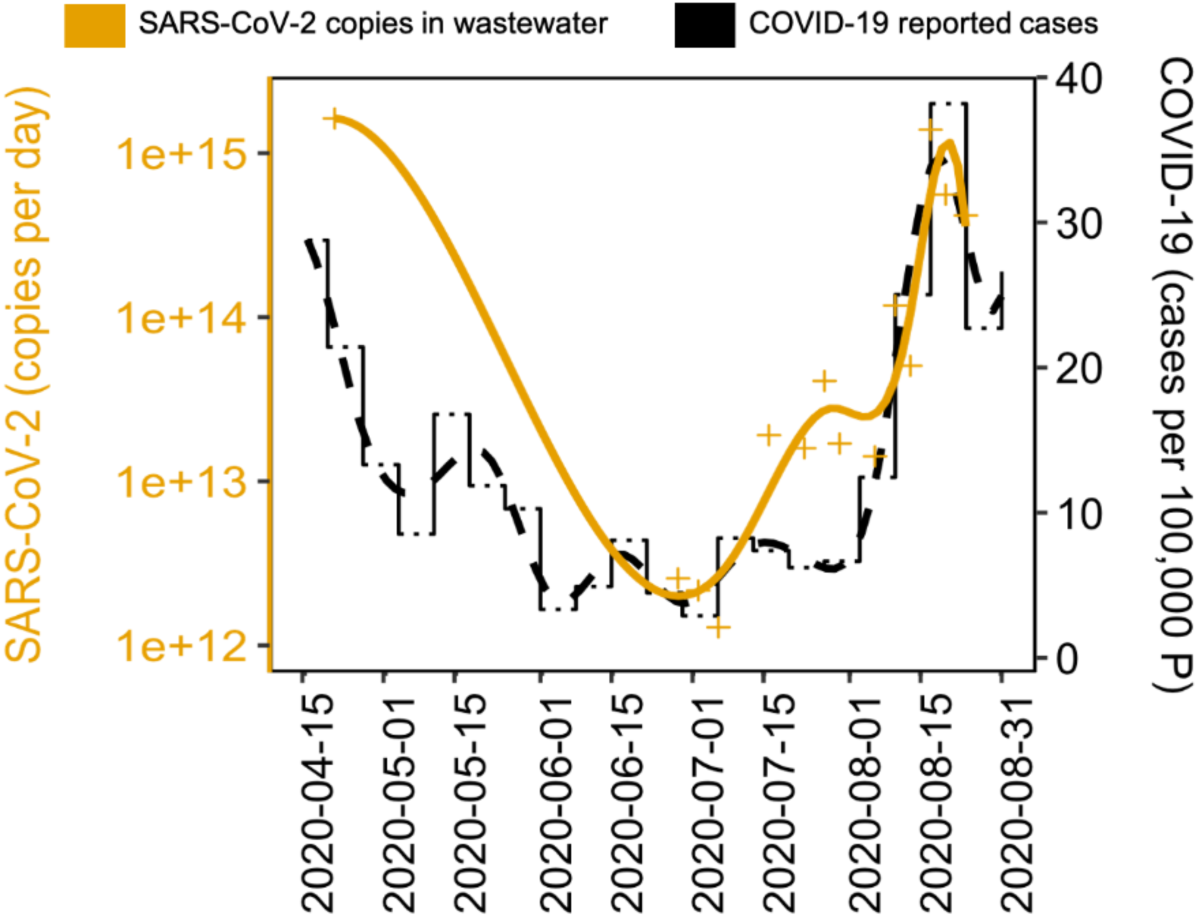
SARS-CoV-2 load as sum of the two WWTP influents as analyzed with RT-qPCR in comparison to the positive tested COVID-19 cases in the city of Frankfurt am Main.

### Multiple WWTPs in the same metropolitan area

The Frankfurt metropolitan area has two main WWTPs, each receiving wastewater from different parts of the city and some neighboring communities. We monitored the influent of both the WWTPs to determine whether there is a significant difference in the viral load in the sewage or its dynamic. After the first peak period of reported positive cases during March and April (Figure 1), the daily load of SARS-CoV-2 to the WWTP Niederrad indicated a slow increase of SARS-CoV-2 over time, with two distinct peaks on the 13^th^ of July (i.e. 6.16 x10^14^ copies/day) and the 17^th^ of August (i.e. 1.36 x10^15^ copies/day) (Figure 4). The increase in load in August is in line with the increase of COVID-19 cases in the studied area. However, the daily load of SARS-CoV-2 in the influent of the WWTP Sindlingen was lower than the one in the influent of the WWTP Niederrad, remaining very similar until the middle of August with values between 1.20 x10^11^ copies/day and 3.17 x10^12^ copies/day. The values increased by a factor of 10 in August to a maximum value of 3.59 x10^13^ copies/day when the COVID-19 cases also increased (Figure 4). The results of the measurements in the influent to the WWTP Sindlingen also revealed a drop in virus load after the peak value of 3.59 x10^13^ copies/day on the 13^th^ of August 2020. We detected a certain SARS-CoV-2 virus load in all the studied samples during this 2-months period for both WWTP influents, with the highest load in the middle of August. The highest detected load also coincided with the end of the summer vacation season in the State of Hesse.

**Figure 4:**
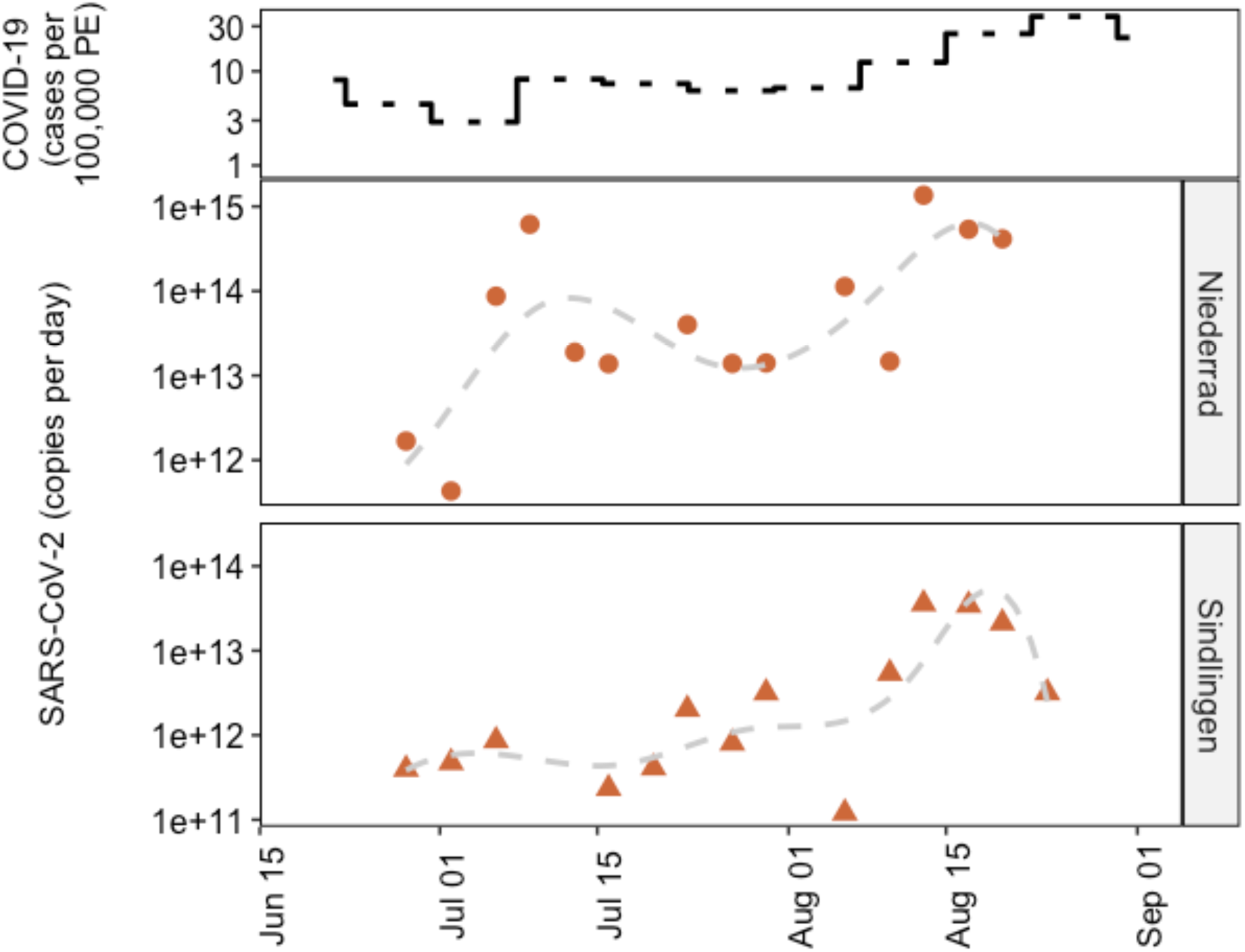
SARS-CoV-2 loads analyzed with RT-qPCR in the influent of the two WWTP, i.e. Niederrad and Sindlingen, and the count of registered COVID-19 cases in the city of Frankfurt (top).

Additionally, to the reported loads, the concentrations of SARS-CoV-2 in the influents to the WWTPs are provided in the Supplementary Material in Figures S.2 and S.3. The concentration of SARS-CoV-2 ranged from 2.0 x10^3^ copies/L to 3.0 x10^6^ copies/L in the influent of the WWTP Niederrad and 3.0 x10^3^ copies/L to 3.0 x10^5^ copies/L in the influent of the WWTP Sindlingen. The concentrations showed very similar trends in comparison the loads. Even though, load based data is always preferred when dealing with wastewater, for SARS-CoV-2 the measured concentrations were able to already provide good approximations of the relative trends given that enough data points are available and no comparison with other samples is required.

The WWTP Niederrad also receives sewage from Griesheim, a part of the Frankfurt metropolitan area, through a separate canal where sampling was possible. Therefore, to generate more localized information, we separately monitored sewage from Griesheim as well. Interestingly, we did not detect any SARS-CoV-2 in a few of the samples from Griesheim during the study period. From the 14 sampling days, six samples showed results of SARS-CoV-2 below the limit of detection (LOD), especially at the beginning of the study in June and July 2020, which corresponds well with the low number of COVID-19 cases reported during this time. Like other sampling points, a moderate increase of virus load in the wastewater from Griesheim was observed, with a specific peak in August when the load was as high as 6.48 x 10^13^ copies/day (see Figure 5).

**Figure 5:**
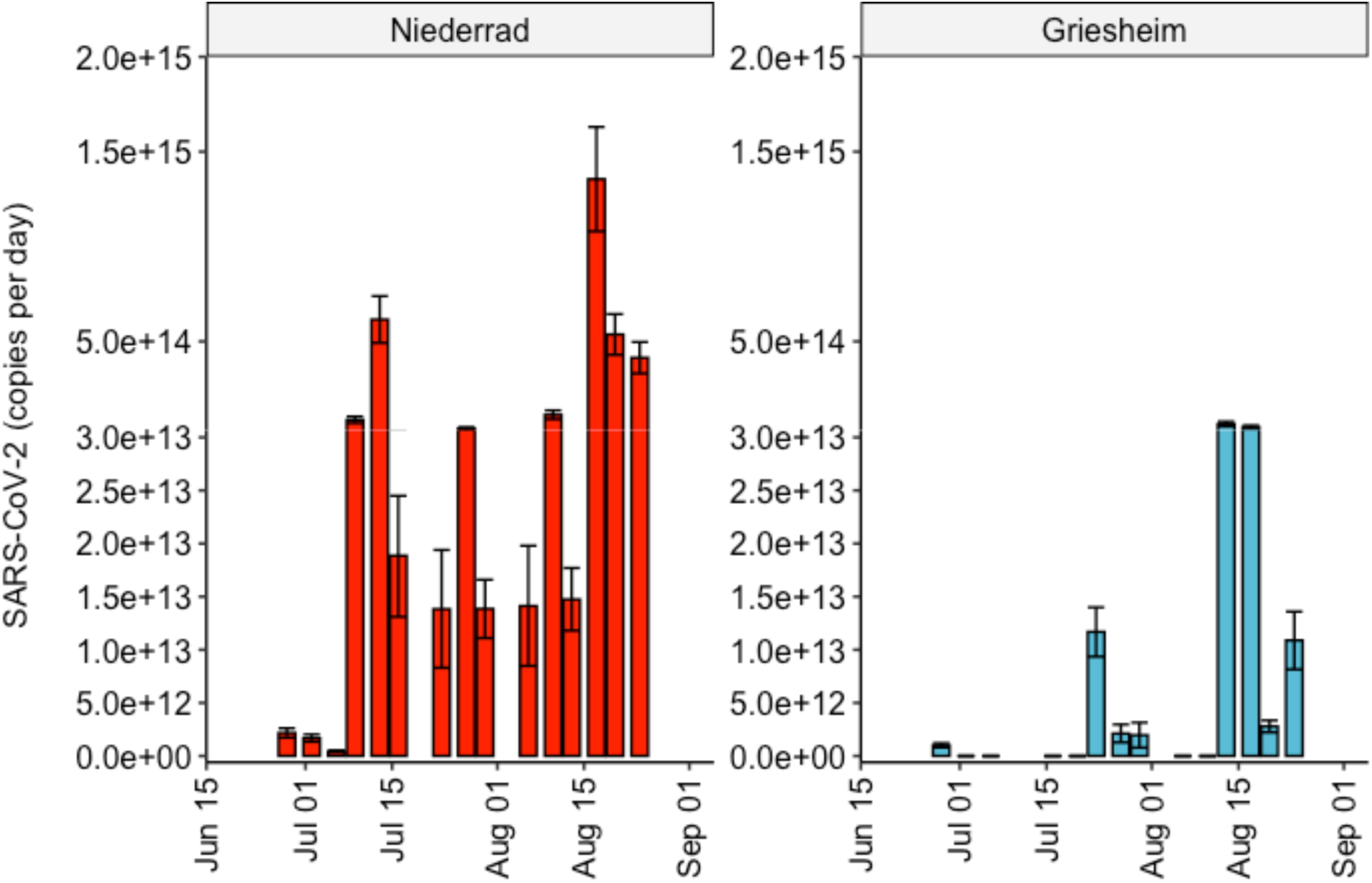
Load of SARS-CoV-2 target genes observed at the sampling point Griesheim and the WWTP Niederrad, respective concentrations of these two sampling points are provided in the supplementary information in Figures S.2 and S.4.

Our results exhibit the epidemiological patterns in the Frankfurt metropolitan area, where WBE indicated an increase of COVID-19 cases through an increase in the virus load to the WWTPs two weeks earlier than the case reports. This result underlines that when cases resurged in August, our WBE analysis could have foreshadowed the increase in positive cases by 10 to 14 days. Similar observations have been reported in other studies ^10,13,14^, showing temporal alignment between the COVID-19 cases and the measured concentrations of the SARS-CoV-2 virus in wastewater. Previous studies of WBE SARS-CoV-2 surveillance in Europe focused on a random sampling of different WWTPs, but similar long-term studies of WBE of SARS-CoV-2 in Europe indicated similar virus concentrations in the wastewater ^10,13^. There is only one known study conducted for German WWTPs ^15^. In this study, the authors measured the SARS-CoV-2 concentration in nine WWTPs in western Germany and reported a load of 1 x 10^11^ to 1 x 10^13^ SARS-CoV-2 copies/day in each of these WWTPs. However, this study reports only one time-point sample.

It is important to note that previous studies vary not only in the sample volume, but also in the protocol of concentration of the virus and as well as the chosen target genes for analysis, which also comprises the use of different primer sets. For this reason, a comparison of SARS-CoV-2 virus loads and concentrations from other WWTPs with the ones reported in this study is not recommended, and preferably, the focus should lie on evaluating the trend of the virus load over time as the more significant and striking information.

Due to the rapid development of the pandemic and the need for testing, several companies have designed qRT-PCR assays for the detection of SARS-CoV-2, but the performance of these assays may vary, esp. since they are designed for medical diagnostics and not for the use with environmental samples such as wastewaters ^16,17^. The results for the different target genes used in this study underlines (Supplementary Figure) the necessity to analyze more than one target gene for better confidence and reliable results. Other studies have also reported variation in the results due to choose target genes and primers ^10,12,15^.

Previously, studies have reported that wastewater with higher density of particles generally yielded lower virus loads, especially with focus on enveloped viruses as they have greater adsorption capabilities to the solid fraction in the wastewater ^18,19^. Therefore, it is important to estimate the viral recovery efficiency, as also suggested in other studies ^17,20^. In our work, wastewater samples were spiked with MS2 phages as a positive control and the recovery efficiency varied between 12 to 89% without significant differences between the recovery efficiency across the different sampling points. Lu et al. (2020) investigated various methods for concentration of the viral RNA in relation to the recovery efficiency. In general, Enterobacteria phages T3 and MS2 demonstrated higher mean recoveries compared to murine hepatitis viruses, spiked porcine epidemic diarrhea viruses and mengoviruses ^21^. The recovery rates reported in the literature varied between 0-21.4 % ^22^ as lowest for SARS-CoV-1 and 85.5 ± 24.5 for Enterobacteria phages MS2 ^19^.

Our study showed, that WBE with the detection of SARS-CoV-2 is a promising method for monitoring local developments. However, there is still the need for more long-term studies to understand and improve the sensitive steps of the analytical methods and gain more knowledge about the reliability of the trends in wastewater samples. The investigated samples in this study included almost exclusively dry weather samples, as the summer month July and August 2020 in Germany were hot and dry without heavy rain events. In future studies, it is thus also necessary to further consider the impact of sewer retention times, stormwater runoff events and the influence of wash-out on SARS-CoV-2 virus loads.

## Data Availability

The data related to the manuscript is provided in the supplementary information.

## Acknowledgment

This work was funded by the Hessian Ministry of Economics, Energy, Transport and Housing within the framework of the IWB-EFRE program for knowledge and technology transfer. We would also like to thank the employees of the Stadtentwässerung Frankfurt (SEF) for their support with sample collection.

## Supplementary Information

### Methods

#### qPCR analysis

We performed qPCR analysis using the Taq Path COVID-19 RT-PCR Kit (Thermo Fisher Scientific) which includes^1^: (1) TaqPath™ COVID-19 Assay Multiplex, which contain three primer/probe sets specific to different SARS-CoV-2 genomic regions (i.e. N gene, S gene and Orf1ab gene) and primers/probes for bacteriophage MS2. (2) MS2 Phage Control – RNA control, having a concentration of 10^6^ copies per µl, to verify the efficacy of the RNA extraction and the absence of inhibitors in the PCR reaction. (3) TaqPath™ COVID-19 Control – Positive SARS-COV2 RNA control that contains targets specific to the SARS-CoV-2 genomic regions targeted by the assays. The manufacturer (Thermo Fisher Scientific) has not publicly released the primers/probe sets sequences, therefore, we do not have access to the information related the primers/probe sets sequences and the length of the PCR products.

In case of the positive control, we included for each qPCR run triplicates of the four different concentration (i.e. 1×10^1^, 2×10^1^, 2×10^2^, 2×10^3^ copies per reaction) of the TaqPath™ COVID-19 positive control. For MS2 phage positive control in each qPCR run triplicates of the three different concentration (i.e. 2×10^2^, 2×10^3^, 2×10^4^ copies per reaction) were also included. For SARS-Cov-2 positive control and MS2 phage positive control, each reaction contained 12.5 µL TaqPath™ 1-Step Multiplex Master Mix (4X), 2.5 µL COVID-19 Real Time PCR Assay Multiplex, 33 µL nuclease free water, and 2 µL of positive control. Triplicate of negative controls were also included in each run, each reaction contained 12.5 µL TaqPath™ 1-Step Multiplex Master Mix (4X), 2.5 µL COVID-19 Real Time PCR Assay Multiplex, and 35 µL nuclease free water. Ct values of positive control dilutions were plotted against known concentration of the SARS-CoV-2 positive control and MS2 phage positive control, to generate standard curves. Start baseline value was set at 5 and Threshold cycle (Ct) values were determined manually while adjusting the threshold to be above any background signal and within exponential phase of the fluorescence curves. Primer efficiencies were 95.32 ± 9.09% for 91.09 ± 13.84% for S, 86.75 ± 1.8% for Orf1ab, and 95.92 ± 17.16%for MS2 phage (n = 8 runs, mean ± sd).

### Results

#### Recovery efficiency of the MS2 phages results

The recovery efficiency of the concentration and extraction procedure performed in triplicates, was determined by using the non-enveloped *Enterobacteria* MS2 phage. It showed an average recovery in the range of 11.53 - 89.11 %, with a median value of 42.40%.

#### Impact of chosen target genes

S.figure 1 shows that different target genes performed differently, especially until the middle of July, when less COVID-19 positive cases were reported. Moreover, variation in the performance of target genes differed with different samples. For example, in initial Niederrad samples, we detected SARS-CoV-2 ORF1ab gene copies only. Whereas, for Sindlingen samples ORF1ab and S gene copies were detected. Based on the results, we recommend targeting multiple genes for SARS-CoV-2 monitoring in wastewater.

#### Concentrations of SARS-CoV-2 in the untreated wastewaters

**S.figure 1:**
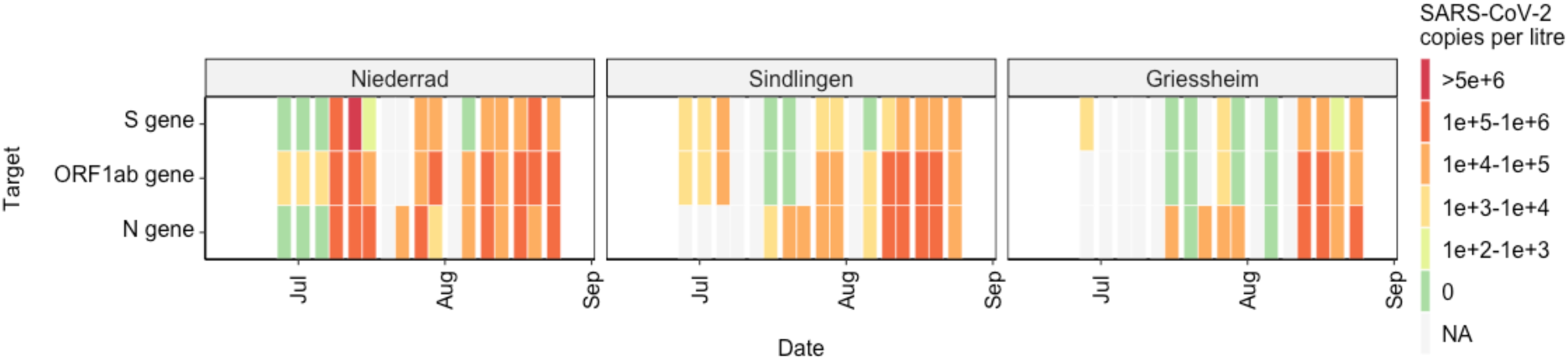
Heatmap showing the SARS-CoV-2 concentration measured for each sample from each sampling point based on three different target genes (S gene, ORF1ab gene, and N gene).NA: Not detected.

**S.figure 2:**
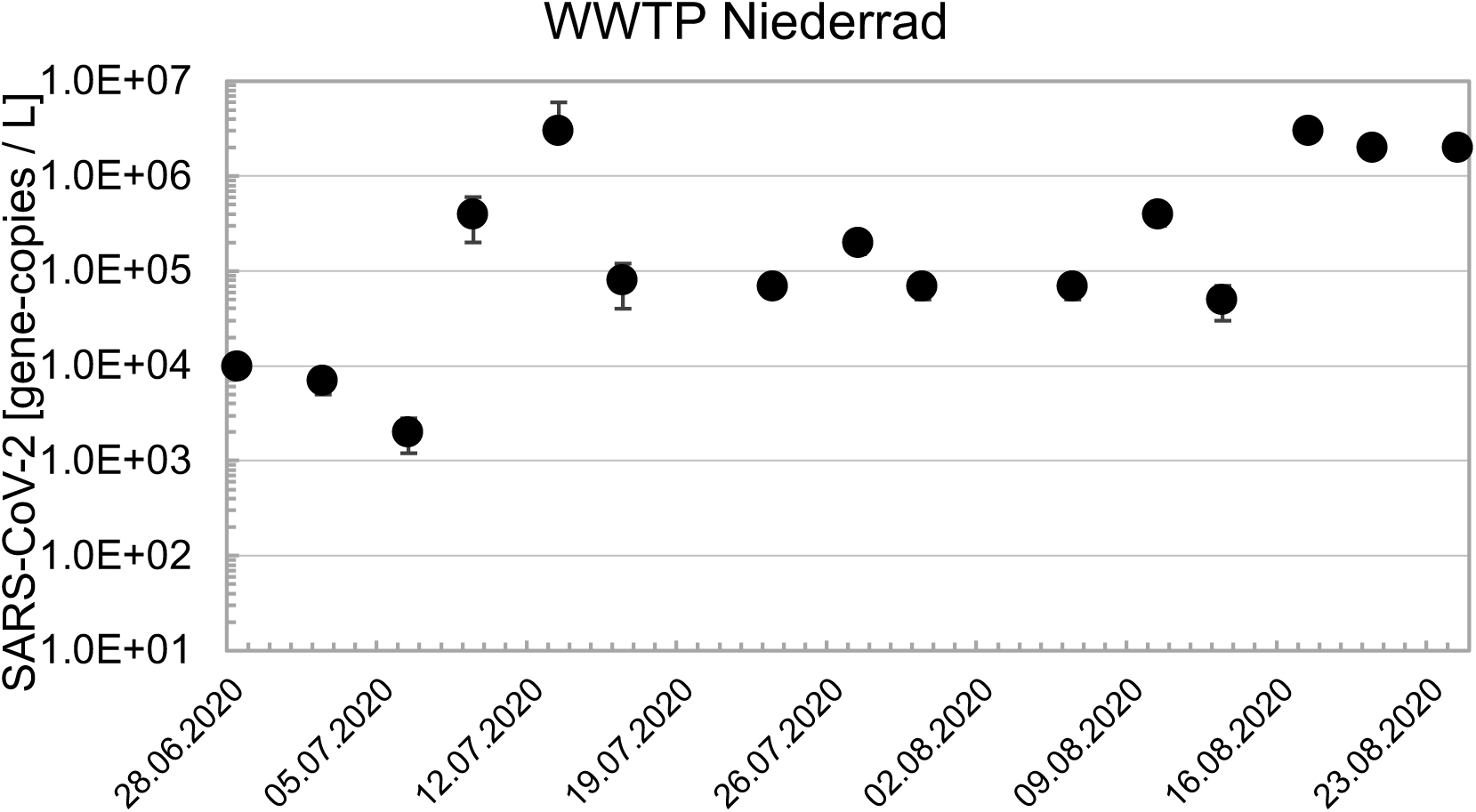
Concentrations of SARS-CoV-2 in the influent of the WWTP Niederrad as determined by real-time qPCR.

**S.figure 3:**
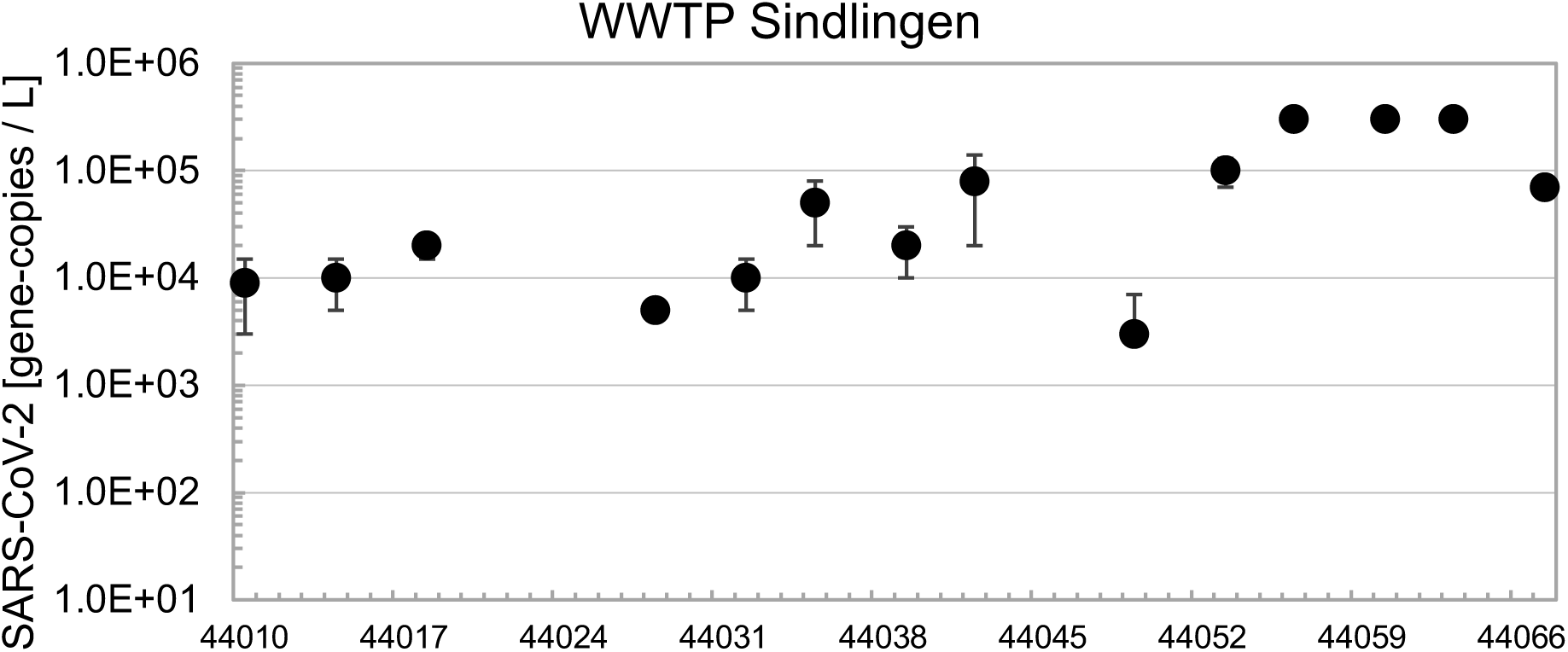
Concentration of SARS-CoV-2 in influent of the WWTP Sindlingen as determined by real-time qPCR.

**S.figure 4:**
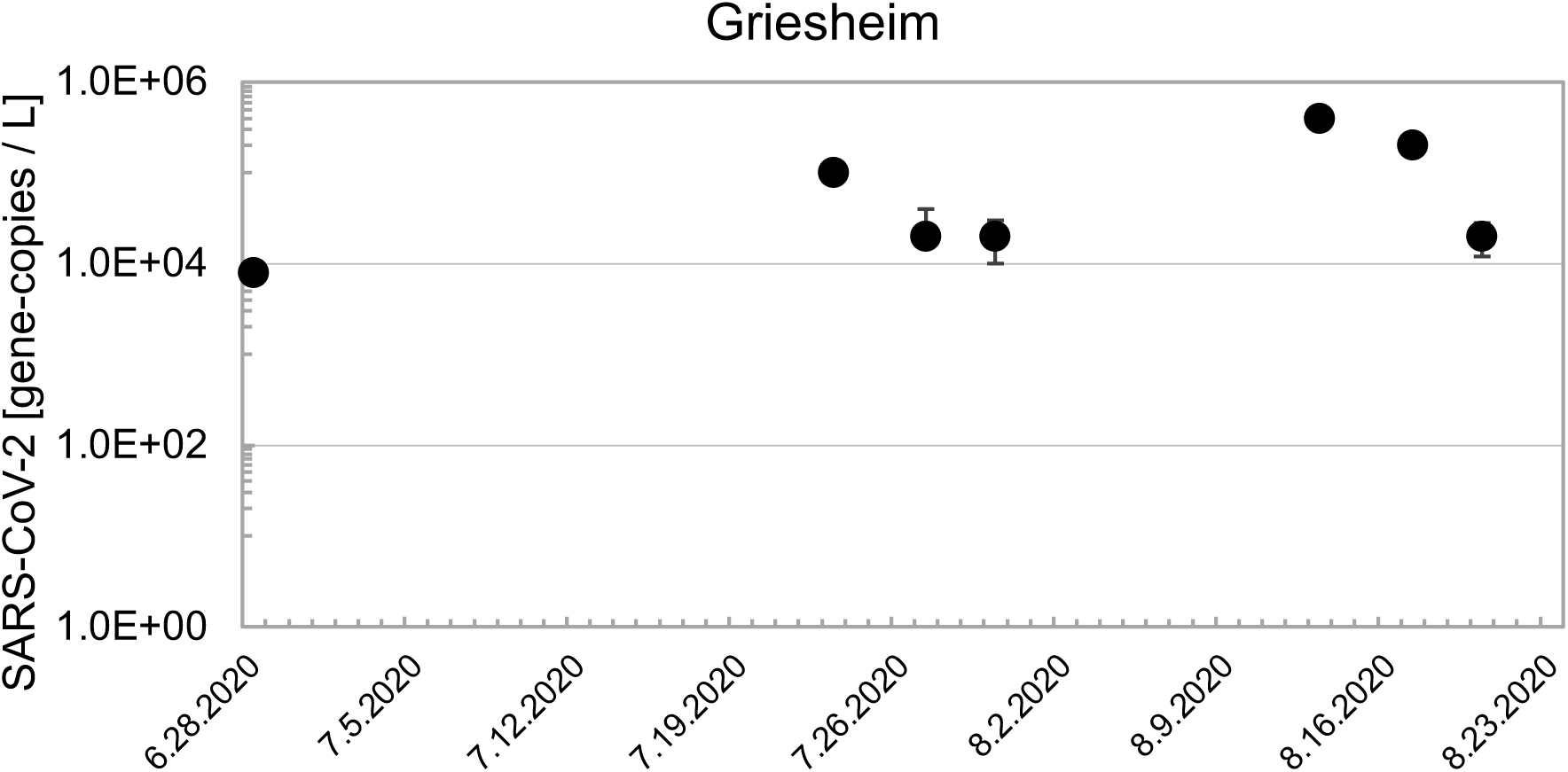
Concentration of SARS-CoV-2 in the wastewater at sampling point Griesheim as determined by real-time qPCR.

## Notes

### Competing Interest Statement

The authors have declared no competing interest.

### Author Declarations

We confirm that all relevant ethical guidelines have been followed.

